# SARS-CoV-2 seroprevalence among healthcare workers in general hospitals and clinics in Japan

**DOI:** 10.1101/2021.01.23.21249922

**Authors:** Tatsuya Yoshihara, Kazuya Ito, Masayoshi Zaitsu, Eunhee Chung, Izumi Aoyagi, Yoshikazu Kaji, Tomomi Tsuru, Takuma Yonemura, Koji Yamaguchi, Shinichi Nakayama, Yosuke Tanaka, Nobuo Yurino, Hideki Koyanagi, Shunji Matsuki, Ryuji Urae, Shin Irie

## Abstract

Coronavirus disease 2019 (COVID-19) has become a serious public health problem worldwide. However, little is known about the prevalence of COVID-19 among healthcare workers in Japan. We aimed to examine the seroprevalence of severe acute respiratory syndrome coronavirus-2 (SARS-CoV-2) antibodies among 2,160 healthcare workers in general hospitals and clinics in Japan. The prevalence of SARS-CoV-2 immunoglobulin G was 1.2% in August and October 2020, which is relatively higher than that in the general population in Japan. Because of the higher risk of COVID-19 infection, healthcare workers should be the top priority for further social support and vaccination against SARS-CoV-2.

## Introduction

Coronavirus disease 2019 (COVID-19) has become a serious public health problem worldwide. In Japan, the confirmed cases of COVID-19 remained low as of December 2020 compared with those in Europe or northern America. The low seroprevalence (0.03%–0.40%) of COVID-19 in the general population of Japan was reported in several studies conducted from June to September 2020 (1). However, the prevalence of COVID-19 among healthcare workers in Japan, who are at higher risk for COVID-19 infection (2), is not well characterized. Here, we aimed to determine the seroprevalence of immunoglobin (Ig)M and IgG antibodies against severe acute respiratory syndrome coronavirus-2 (SARS-CoV-2) among workers in general hospitals and clinics in Japan.

## Methods

A multicenter prospective study was conducted in nine general hospitals and clinics of the SOUSEIKAI Medical Group: Fukuoka Mirai Hospital (FMH), Hakata Clinic (HC), PS Clinic (PC), Sumida Hospital (SH), Miyata Hospital (MH), Kanenokuma Hospital (KH), Shinyoshizuka Hospital (SYH), Nishikumamoto Hospital (NH), and DouDou Clinic (DC). These hospitals/clinics are not designated to treat COVID-19 patients. FMH Clinical Research Center, HC, NH Clinical Pharmacology Center, and SH are specialized facilities for clinical trials (mainly Phase 1 clinical trials). FMH, PC, MH, KH, and SYH are located in Fukuoka prefecture; NH is located in Kumamoto prefecture; and DC and SH are located in Tokyo.

This study was approved by the SOUSEIKAI Hakata Clinic Institutional Review Board (approval number: N-105) and registered in the UMIN Clinical Trial Registry (registration number: UMIN000041262). The people working in the hospitals/clinics of the SOUSEIKAI Medical Group were invited to participate in the study. All participants provided a written informed consent. In each survey, a questionnaire was used to obtain the following data: job title, presence of suspected COVID-19 symptoms since February 2020, and history of COVID-19 diagnosis. The participants’ occupations were divided into eight categories: nurses (including nurse assistants), physicians, technicians (laboratory technicians, radiology technicians, pharmacists, clinical engineers, dental hygienists, physical therapists, occupational therapists, speech therapists, and acupuncturists), nursing care staff, office workers, receptionists, employees in clinical research units, and others (drivers, security personnel, nursery school teachers, shop workers, sanitary workers, nutritionists, and food service staff).

SARS-CoV-2-specific IgM and IgG antibodies in the venous blood were assessed using an immunochromatographic assay kit (2019-nCoV Ab Test [Colloidal Gold], INNOVITA Biological Technology Co., Ltd., Tangshan, China) following the manufacturer’s instructions. Blood samples were collected in August and/or October 2020. Using descriptive statistics, the difference in seropositive rates across background characteristics was assessed using a chi-square test. Two-tailed P values of <0.05 were considered significant. Statistical analyses were performed using JMP Pro 15 (SAS Institute Inc. Japan, Tokyo, Japan).

## Results

A total of 2,160 SOUSEIKAI workers, aged 20–83 years (mean = 41.9, standard deviation =12.7; women: 1,547 [71.6%]), underwent at least one antibody test in August and/or October. IgM and/or IgG SARS-CoV-2 antibodies were detected in 33 (1.5%) and 27 (1.3%) participants in August and October, respectively (Table 1). IgG antibodies against SARS-CoV-2 were detected in 25 (1.2%) and 24 (1.2%) participants in August and October, respectively. After excluding one facility, where nosocomial infections occurred in April 2020, 13 participants (0.8%) demonstrated IgG positivity in August and October (Table 1). Of the eight participants who showed IgM positivity in August, none reported IgG positivity in October. Among the participants who showed IgG positivity in August, 95.5% (21/22) demonstrated IgG positivity in October.

**Table 1.** Prevalence of SARS2-CoV2 antibodies in August or October 2020 stratified by region

| Region (prefecture) | August 2020 |  |  |  | October 2020 |  |  |  |
| --- | --- | --- | --- | --- | --- | --- | --- | --- |
|  | n | Positive, n (%) |  |  | n | Positive, n (%) |  |  |
|  |  | IgM | IgG | IgM and/or IgG |  | IgM | IgG | IgM and/or IgG |
| Fukuoka | 1,746 | 10 (0.6) | 25 (1.4) | 33 (1.9) | 1,698 | 3 (0.2) | 24 (1.4) | 27 (1.6) |
| Kumamoto | 321 | 0 (0) | 0 (0) | 0 (0) | 308 | 0 (0) | 0 (0) | 0 (0) |
| Tokyo | 75 | 0 (0) | 0 (0) | 0 (0) | 75 | 0 (0) | 0 (0) | 0 (0) |
| Total | 2,142 | 10 (0.5) | 25 (1.2) | 33 (1.5) | 2,081 | 3 (0.1) | 24 (1.2) | 27 (1.3) |
| Comparison between the facilities where nosocomial infection occurred and in those where nosocomial infection did not occur |  |  |  |  |  |  |  |  |
| Nosocomial infection (−) | 1,654 | 10 (0.6) | 13 (0.8) | 21 (1.3) | 1,595 | 3 (0.2) | 13 (0.8) | 16 (1.0) |
| Nosocomial infection (+) | 488 | 0 (0) | 12 (2.5) | 12 (2.5) | 486 | 0 (0) | 11 (2.3) | 11 (2.3) |
| P value<sup>a,b</sup> |  | 0.09 | 0.003 | 0.06 |  | 0.34 | 0.009 | 0.03 |

<sup>a</sup>Pearson's chi-square test
<sup>b</sup>Comparison between the facilities where nosocomial infection occurred and those where nosocomial infection did not occur

The prevalence of IgG antibodies was higher among the 20–29-year and 60– 69-year age groups compared with other age groups, whereas no significant difference was observed between women and men (Table 2). The prevalence of IgG antibodies was relatively, but not significantly, higher among nurses, nursing care staff, and receptionists (Table 2). Among the 28 participants who had at least one IgG-positive test result, 17 (60.7%) had suspected symptoms of COVID-19 since February 2020. Among the 14 participants diagnosed with COVID-19 prior to the antibody tests, 10 (71.4%) tested positive for IgG antibodies.

**Table 2.** Prevalence of SARS-CoV-2 antibodies in August and/or October 2020 stratified by COVID-19 history, decade age group, sex, and job categories

| Characteristics | n | IgM Positive<br>n (%) | IgG Positive<br>n (%) | IgM and/or IgG<br>Positive, n (%) |
| --- | --- | --- | --- | --- |
| Total | 2,160 | 10 (0.5) | 28 (1.3) | 36 (1.7) |
| COVID-19 history (–) | 2,146 | 10 (0.5) | 18 (0.8) | 26 (1.2) |
| COVID-19 history (+) | 14 | 0 (0) | 10 (71.4) | 10 (71.4) |
| Age |  |  |  |  |
| 20–29 | 411 | 2 (0.5) | 11 (2.7) | 12 (2.9) |
| 30–39 | 587 | 3 (0.5) | 1 (0.2) | 3 (0.5) |
| 40–49 | 601 | 3 (0.5) | 7 (1.2) | 10 (1.7) |
| 50–59 | 330 | 0 (0.0) | 4 (1.2) | 4 (1.2) |
| 60–69 | 174 | 2 (1.2) | 5 (2.9) | 7 (4.0) |
| ≥70 | 57 | 0 (0.0) | 0 (0.0) | 0 (0.0) |
| P value<sup>d</sup> |  | 0.60 | 0.006 | 0.007 |
| Sex |  |  |  |  |
| Female | 1,547 | 6 (0.4) | 21 (1.4) | 27 (1.8) |
| Male | 613 | 4 (0.7) | 7 (1.1) | 9 (1.5) |
| P value<sup>d</sup> |  | 0.41 | 0.69 | 0.65 |
| Job categories |  |  |  |  |
| Nurse<sup>a</sup> | 651 | 3 (0.5) | 11 (1.7) | 14 (2.2) |
| Physician | 97 | 0 (0.0) | 1 (1.0) | 1 (1.0) |
| Technician<sup>b</sup> | 369 | 2 (0.5) | 3 (0.8) | 4 (1.1) |
| Nursing care staff | 329 | 2 (0.6) | 8 (2.4) | 9 (2.7) |
| Office worker | 270 | 2 (0.7) | 2 (0.7) | 4 (1.5) |
| Receptionist | 33 | 0 (0.0) | 1 (3.0) | 1 (3.0) |
| Clinical research unit worker | 192 | 0 (0.0) | 0 (0.0) | 0 (0.0) |
| Other<sup>c</sup> | 219 | 1 (0.5) | 2 (0.9) | 3 (1.4) |
| P value<sup>d</sup> |  | 0.95 | 0.25 | 0.33 |

<sup>a</sup>Nurses include assistants
<sup>b</sup>Laboratory technicians, radiology technicians, pharmacists, clinical engineers, dental hygienists, physical therapists, occupational therapists, speech therapists, and acupuncturists <sup>c</sup>Drivers, security personnel, nursery school teachers, workers in shop, sanitary workers, nutritionists, and food service staff <sup>d</sup>Pearson's chi-square test

## Discussion

The prevalence of SARS-CoV-2 antibodies varies among countries and cities. Europe and northern America have high prevalence of SARS-CoV-2 antibodies, whereas Eastern Asia has a relatively low prevalence (3). In Japan, several studies conducted from June to September 2020 reported low seroprevalence of SARS-CoV-2 antibodies (0.03%–0.40%) in the general population (1). In this study, the seroprevalence of SAR-CoV-2 IgG antibodies among healthcare workers was 1.2% in August and October 2020. It has been reported that the prevalence of SARS-CoV-2 antibodies in healthcare workers was higher than that in the general population (2,4). Our study also demonstrated that the prevalence of SARS-CoV-2 antibodies was relatively higher in healthcare workers than in the general population in Japan.

The 20–29-year and 60–69-year age groups had high prevalence of SARS-CoV-2 IgG antibodies. Among the five participants in the 60–69-year age group who tested positive for IgG antibodies, four worked in the facility where nosocomial infection occurred in April 2020; therefore, the high prevalence among the 60–69-year age group was possibly due to nosocomial infection. The routes of infection in the 20–29-year age group have not been fully elucidated. However, the high occupational and daily activities and the demographic characteristics of asymptomatic carriers in the young age group were considered to be contributing factors.

The level of SARS-CoV-2 antibodies remain high for at least a few months after developing a COVID-19 infection (5–7). A recent study reported high levels of SARS-CoV-2 antibodies (69.0%–91.4%) 8 months after asymptomatic or mild SARS-CoV-2 infection (8). In this study, 95% (21/22) of participants who had IgG antibodies in August tested positive for IgG antibodies in October. Among the participants who had a history of acquiring COVID-19, 71.4% (10/14) showed IgG positivity up to 6 months after infection. These results support those of previous studies, which reported that IgG antibodies can be detected in the majority of patients a few months after SARS-CoV-2 infection.

One limitation of this study is that only one antibody assay kit was used. As PCR tests or antigen tests were not conducted in participants with SARS-CoV-2 IgM or IgG antibodies, it remained uncertain whether false-positive data were included. In particular, among the eight participants who tested positive for IgM antibodies alone in August, none tested positive for IgG antibodies in October. We recruited individuals working at SOUSEIKAI medical group facilities for this study; of them, 91.6% (2,142 out of 2,338) in August and 90.0% (2,081 out of 2311) in October participated in this study. Thus, the effect of selection bias is limited.

In conclusion, the prevalence of SARS-CoV-2 IgG antibodies among healthcare workers in Japan was 1.2%, which was relatively higher than that in Japan’s general population. Healthcare workers are at higher risk of infection; hence, they should be the top priority for further social support and SARS-CoV-2 vaccination.

## Data Availability

The data that support the findings of this study are available from the corresponding author, TY, upon reasonable request, according to the Ethical Guidelines for Medical and Health Research Involving Human Subjects, Japan.

## Acknowledgement

The authors would like to thank SoftBank Group Corp. for providing the immunochromatographic assay kits, the workers at the study settings for their consent to participate and support in conducting this study, Ms. Junko Manabe and Yoshie Moriji for collecting the study data, and Editage (www.editage.com) for providing English language editing assistance.

## Conflict of interest

None to declare.

## Funding

This study did not receive external funding. The immunochromatographic assay kits used in this study were provided by SoftBank Group Corp.

## Author contributions

TYoshihara, RU, and SI conceived and designed the study. TY, KI, and SI wrote the protocol of the study. IA, RK, TT, TYonemura, KY, SN, YT, NY, and HK conducted the study in each study setting. TY, KI, MZ, and SI analyzed and interpreted the data. TY, KI, MZ, and EC drafted the manuscript. All authors interpreted the results of the analyses, critically reviewed and edited the manuscript, and approved the final version.

## Notes

### Competing Interest Statement

The authors have declared no competing interest.

### Clinical Trial

UMIN000041262

### Author Declarations

SOUSEIKAI Hakata Clinic Institutional Review Board (Approval number: N-105)

